# Nonlinear Mixed Effect Modeling of Patient-Reported Outcomes to Improve Treatment Effect Estimation in an Oncology Clinical Trial

**DOI:** 10.64898/2026.10.26.26366496

**Authors:** Congyu Zhang, Quefeng Li, Ghassan Slim, Xihao Li, Yangbo Sun, Benyam Muluneh, Jiawei Zhou

## Abstract

**Background:** Patient-reported outcomes (PROs) capture how treatments affect patients’ symptoms, functioning, and quality of life, but substantial variability can obscure treatment effects. We evaluated whether NLME modeling could improve treatment-effect estimation for PRO endpoints by accounting for data variability.

**Methods:** We retrospectively analyzed participant-level data from the phase III coBRIM trial (NCT01689519), comparing cobimetinib plus vemurafenib with placebo plus vemurafenib in melanoma. EORTC QLQ-C30 global health status/quality of life (GHS/QoL) scores were evaluated using observed-score comparisons, mixed models for repeated measures (MMRM), and NLME modeling. The NLME framework, informed by pharmacology knowledge and context, characterized longitudinal GHS/QoL trajectories while quantifying between- and within-patient variability. Model-estimated scores and trajectory parameters were compared between treatments.

**Results:** A total of 202 participants with 1,443 GHS/QoL assessments were analyzed. Observed GHS/QoL scores were highly variable, and neither observed-score comparisons nor MMRM identified statistically significant treatment differences at Cycle 8 Day 1. NLME modeling identified significantly less deterioration in longitudinal GHS/QoL with cobimetinib plus vemurafenib (P = 0.036). Model-estimated GHS/QoL scores were also significantly higher with combination treatment at Cycle 8 Day 1 (P = 0.049). Model simulations estimated a 10.4-point between-group difference (95% interval, −6.2 to 26.9), exceeding a commonly used threshold for clinical importance, although uncertainty remained.

**Conclusions:** NLME modeling can improve PRO data analysis by accounting for substantial between- and within-patient variability and incorporating pharmacological principles to characterize disease- and treatment-related PRO changes over time. This approach may improve treatment-effect estimation and reveal clinically meaningful differences overlooked by conventional analyses.

## Introduction

Cancer represents a substantial global health burden, with an estimated 20.6 million new cases diagnosed worldwide in 2024. Approximately one in five individuals will develop cancer during their lifetime, underscoring the broad population impact of the disease [1]. For patients with cancer, treatment benefit extends beyond tumor response and survival to how they feel and function during therapy [2; 3]. The U.S. Food and Drug Administration has increasingly emphasized patient-focused outcome assessment in oncology, including systematic evaluation of patient-reported symptoms, functioning, and other aspects of health-related quality of life in clinical trials [4; 5]. Patient-reported outcomes (PROs) therefore provide an important complement to conventional efficacy endpoints by directly capturing the patient experience of treatment [6].

Although PROs are increasingly collected in oncology trials, they are most often evaluated as secondary or exploratory endpoints and remain infrequently incorporated into regulatory labeling and formal treatment-benefit claims [7; 8]. One challenge is that treatment effects on PROs can be difficult to interpret because patient-reported data often show substantial variability both between patients and within the same patient over time [9]. Patients differ substantially in how they experience and report the same disease and treatment, resulting in high between-patient variability [10–12]. PRO scores can also fluctuate within the same patient because of sleep, mood, stress, and other day-to-day factors unrelated to sustained clinical change [13; 14]. For example, a patient may report greater fatigue or worse mood on a particular day after a poor night’s sleep, reflecting a short-term fluctuation rather than a sustained change in treatment effect [15]. When data variability is large, comparisons of observed PRO scores at individual study visits may produce imprecise estimates of treatment effects and make meaningful differences in longitudinal trajectories difficult to distinguish from background variability [9]. Consequently, PRO findings may sometimes appear discordant with treatment benefits observed in other clinical outcomes [16; 17]. This pattern has been observed across oncology trials: a meta-epidemiologic analysis of 791 phase 3 studies found improvements in overall survival in 28% of trials, compared with improvements in QoL in only 11%, while only 6% demonstrated improvement in both outcomes [18].

Nonlinear mixed-effects (NLME) modeling offers a pharmacology-informed approach to analyzing longitudinal PRO data by capturing underlying response trajectories and accounting for variability within and between patients. [9]. Widely used in pharmacokinetic and pharmacodynamic analyses, NLME models can incorporate pharmacology-informed nonlinear functions that describe the magnitude and time course of treatment-related changes [19]. When applied to PROs, these trajectories may reflect the combined effects of disease progression, therapeutic efficacy, and treatment-related toxicity on patients’ health status. Moreover, NLME models use fixed effects to characterize population-level trajectories and random effects to quantify between-patient variability, while residual error models account for within-patient variability unexplained by the underlying trajectories. [19] By integrating repeated measurements and separating systematic changes from variability, NLME modeling may provide a more precise characterization of treatment effects than comparisons of observed PRO scores at individual study visits [20].

The phase III coBRIM trial (NCT01689519) provides an opportunity to evaluate this approach [21]. In the coBRIM study, cobimetinib plus vemurafenib improved progression-free survival and overall survival compared with placebo plus vemurafenib, whereas conventional analyses of EORTC QLQ-C30 global health status/quality of life (GHS/QoL) scores showed generally comparable quality of life between treatment groups [21; 22]. We hypothesized that substantial between-patient and within-patient variability may have obscured treatment-related differences in the observed PRO data. In this retrospective analysis, we applied NLME modeling to longitudinal EORTC QLQ-C30 GHS/QoL data to determine whether accounting for these sources of variability and incorporating pharmacology-informed nonlinear functions could more precisely characterize the treatment effect on PROs.

## Methods

### Data

A retrospective, secondary data analysis using de-identified individual participant data from the coBRIM trial (ClinicalTrials.gov Identifier: NCT01689519) was performed in this study. The coBRIM trial was a double-blind, placebo-controlled, randomized phase 3 trial that enrolled adults with previously untreated, unresectable stage IIIC or stage IV melanoma harboring a BRAF^V600^ mutation [21]. Participants were randomly assigned 1:1 to receive cobimetinib plus vemurafenib or placebo plus vemurafenib. Cobimetinib, a selective inhibitor of MEK1 and MEK2 in the MAPK signaling pathway [23], was administered at 60 mg once daily for 21 days followed by 7 days off treatment in each 28-day cycle, and vemurafenib was administered at 960 mg twice daily. In this analysis, 202 patients from the coBRIM trial were available in the sponsor-provided dataset. The analytic dataset included baseline characteristics, treatment assignment, longitudinal PROs, and overall survival data.

### PRO Measure

The primary outcomes for this secondary analysis were longitudinal patient-reported health-related quality of life outcomes assessed with the European Organization for Research and Treatment of Cancer Quality of Life Questionnaire Core 30 (EORTC QLQ-C30) [24]. In the coBRIM trial, the EORTC QLQ-C30 survey was administered at baseline, days 1 and 15 of cycles 1 and 2, and day 1 of every other 28-day cycle thereafter until study withdrawal or the end of study. The EORTC QLQ-C30 includes 2 items specifically assessing GHS/QoL. Question 29 asks patients to rate their overall health during the past week, and question 30 asks them to rate their overall quality of life during the past week, each on a 7-point scale ranging from 1 (very poor) to 7 (excellent). The 2 item scores were averaged and linearly transformed to a 0- to 100-point scale according to the EORTC scoring procedure, with higher scores indicating better GHS/QoL [25]. In this analysis, the responses to Questions 29 and 30 were used as the longitudinal GHS/QoL outcome for this secondary analysis.

### NLME Modeling Approach

NLME modeling approach incorporating item response theory (IRT) was used to analyze GHS/QoL data. [26] Rather than comparing GHS/QoL scores separately at each study visit, the NLME modeling approach describes the pattern of PROs over time, allowing the changes in PROs to be nonlinear and influenced by both disease and treatment. [9] The longitudinal GHS/QoL trajectory was described using a sigmoidal function commonly applied in pharmacodynamic modeling. [27] This function characterizes the magnitude (*Emax*) and rate (*Kd*) of change in GHS/QoL over time, providing a pharmacologically motivated representation of the evolving effects of disease and treatment on patient-reported health status (**Equation 1**).

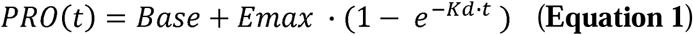

Model parameters were estimated using data from all patients to characterize the average PRO trajectory in the cohort while allowing each patient to have a unique trajectory (**Equation 2**). This approach can quantify differences in baseline PRO scores and variability in PRO trajectories between patients, as well as variability within individual patients (**Equation 3**).

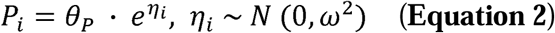

Where *P_i_* represents the individual parameter value for subject *i*, *θ_P_* is the population typical value, and *η_i_* represents between-patient variability.

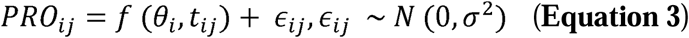

Where *PRO_i,j_* is the observed GHS/QoL scores in subject *i* at time *j*. *f*(*θ_i_*, *t_ij_*) represents the underlying longitudinal GHS/QoL trajectory for subject *i* at time *j*, and *∈_ij_* represents residual unexplained variability.

By accounting for these sources of variability, this model can separate disease- or treatment-associated changes in PROs from random or short-term fluctuations that may not reflect clinically meaningful change. Model-predicted GHS/QoL scores and trajectory parameters therefore represent the underlying longitudinal PRO trajectory after excluding residual within-patient variability. Additional details of the NLME modeling approach are provided in **Supplementary Methods**.

## Statistical Analysis

Statistical analyses compared treatment effects on PRO endpoints using observed and NLME model-based approaches. Observed GHS/QoL scores at Cycle 8 Day 1 were compared between treatment arms using Wilcoxon rank-sum tests. A mixed model for repeated measures (MMRM) was also used to estimate least-squares mean changes from baseline in GHS/QoL scores and between-group treatment differences at each post-baseline visit. For the NLME modeling approach, model-derived patient-specific PRO trajectory parameters and model-predicted GHS/QoL scores at Cycle 8 Day 1 were compared between treatment arms using Wilcoxon rank-sum tests. To evaluate the clinical relevance of the NLME modeling-predicted PRO trajectories, adjusted multivariable Cox proportional hazards models were used to assess associations between PRO trajectory parameters and survival outcomes. Two-sided P values <0.05 were considered statistically significant.

## Software and Environment

All analyses were conducted within the Vivli research environment. Data cleaning, statistical analyses and data visualization were performed using R 4.4.0 and RStudio Version 2024.12.0. NLME model estimation was performed using Monolix 2024R1 (Lixoft), and model simulations were performed using Simulx 2024R1. Both Monolix and Simulx could be downloaded at https://lixoft.com/products/. The figures were compiled in Adobe Illustrator 2025. The analysis code is available on GitHub (https://github.com/zhangcy000/CoBRIM-Trial-Code)

## Data Availability

The source data used in this study were provided through the Vivli data-sharing platform. Access to the data is subject to Vivli’s data access procedures and applicable data use agreements.

## Results

### Patient Characteristics

A total of 202 participants with 1,443 PRO assessments were included in this secondary analysis. The median age was 55.5 years (range, 23 - 86 years); 77 (38%) were female and 125 (62%) were male. 194 participants (96%) were White, and 8 (4%) were of other races. 103 participants (51%) were randomized to receive cobimetinib plus vemurafenib and 99 (49%) to receive placebo plus vemurafenib. Baseline patient characteristics are summarized in **Table 1**.

**Table 1.** Baseline demographic and clinical characteristics.

|  | **Cobimetinib +**<br>**Vemurafenib**<br>**(N = 103)** | **Placebo +**<br>**Vemurafenib**<br>**(N = 99)** | **Total**<br>**(N = 202)** |
| --- | --- | --- | --- |
| **AGE, median (range),** | 55 (23, 86) | 56 (25, 80) | 55.5 (23, 86) |
| **SEX** |  |  |  |
| Male | 66 (64%) | 59 (60%) | 125 (62%) |
| Female | 37 (36%) | 40 (40%) | 77 (38%) |
| **Body weight, median** | 83.2 (50.5 – 185) | 83 (46.2 – 148.1) | 83 (46.2, 185) |
| **Race** |  |  |  |
| White | 99 (96%) | 95 (96%) | 194 (96%) |
| Other | 4 (4%) | 4 (4%) | 8 (4%) |
| **Geographical region** |  |  |  |
| Asia | 16 (16%) | 16 (16%) | 32 (16%) |
| Europe | 29 (28%) | 26 (26%) | 55 (27%) |
| North America | 25 (24%) | 26 (26%) | 51 (25%) |
| Oceania | 33 (32%) | 31 (31%) | 64 (32%) |
| **ECOG performance** |  |  |  |
| 0 | 75 (73%) | 62 (63%) | 137 (68%) |
| 1 | 26 (25%) | 37 (37%) | 63 (31%) |
| Unknown | 2 (2%) | 0 | 2 (1%) |
| **Metastatic Status** |  |  |  |
| Unresectable stage IIIC | 5 (4.9%) | 2 (2%) | 7 (4%) |
| M1a | 17 (17%) | 15 (15%) | 32 (16%) |
| M1b | 19 (18%) | 18 (18%) | 37 (18%) |
| M1c | 62 (60%) | 64 (65%) | 126 (62%) |
| **BRAF mutation** |  |  |  |
| V600E | 69 (67%) | 69 (70%) | 138 (68%) |
| V600K | 7 (6.8%) | 13 (13%) | 20 (10%) |
| Not done | 27 (26%) | 17 (17%) | 44 (22%) |

| **Previous adjuvant** |  |  |  |
| --- | --- | --- | --- |
| Yes | 9 (8.7%) | 7 (7.1%) | 16 (8%) |
| No | 94 (91%) | 92 (93%) | 186 (92%) |
| **Lactate dehydrogenase** |  |  |  |
| Raised | 39 (38%) | 39 (39%) | 78 (39%) |
| Normal | 64 (62%) | 60 (61%) | 124 (61%) |

### Observed GHS/QoL Scores

Observed GHS/QoL scores between the cobimetinib plus vemurafenib and vemurafenib groups throughout the evaluable period were reported in **Figure 1**. At Cycle 8 Day 1, the mean (SD) change from baseline was -0.44 (17.28) points in the cobimetinib plus vemurafenib group and - 7.72 (23.0) points in the vemurafenib group. Although the vemurafenib group had a numerically greater mean decline in quality of life, substantial within-group variability was observed, and the between-group difference was not statistically significant (P = 0.09).

**Figure 1.**
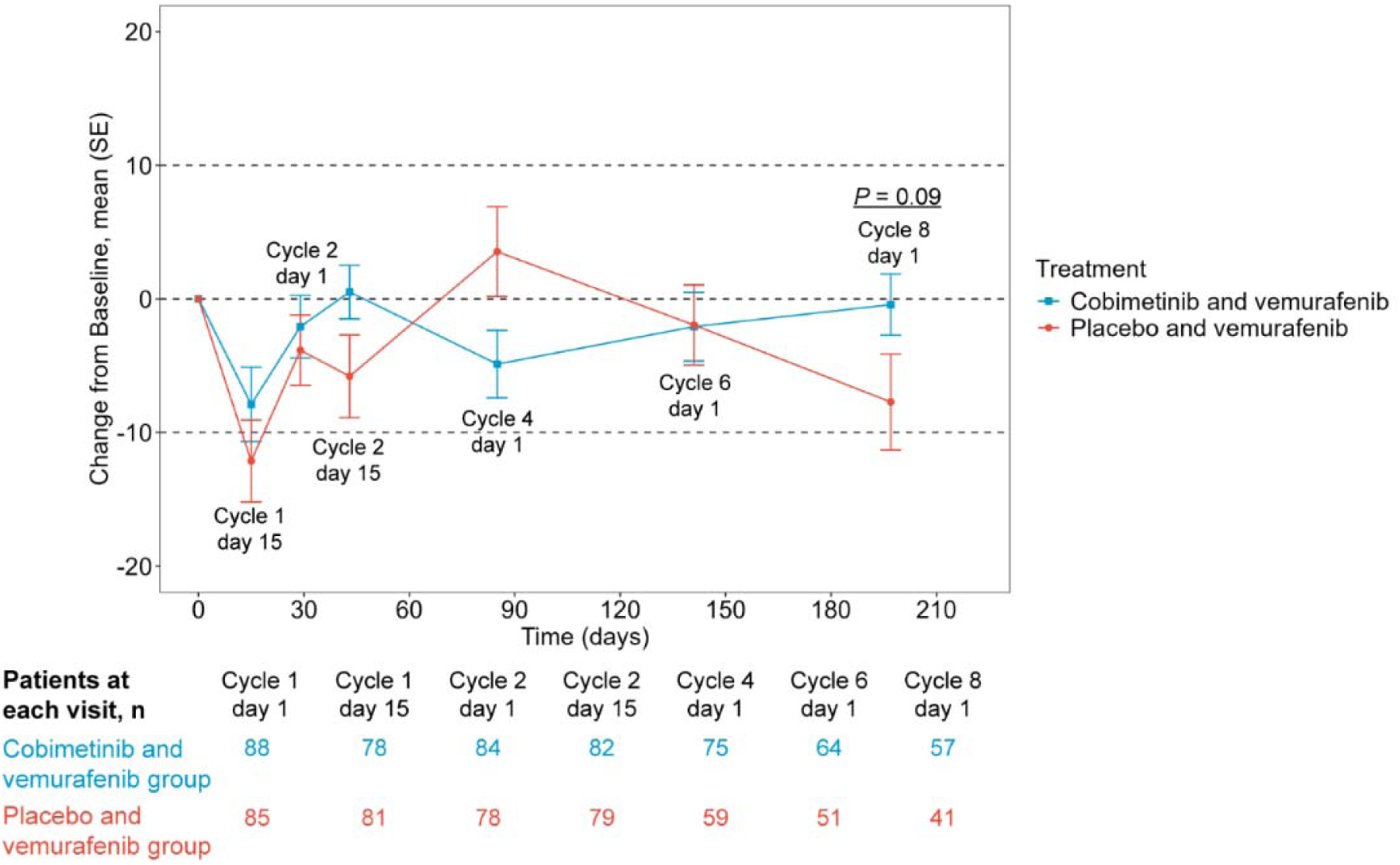
Observed mean change from baseline in EORTC QLQ-C30 GHS/QoL scores. The dashed horizontal lines at ±10 points represent the threshold for clinically meaningful improvement or deterioration from baseline in global health status. The numbers of evaluable patients at each assessment are shown below the figure.

Similarly, MMRM did not identify statistically significant between-group differences in GHS/QoL scores change from baseline across study visits as shown in **Table S1**. At Cycle 8 Day 1, the least-squares mean change from baseline was −2.84 points with cobimetinib plus vemurafenib and −7.52 points with placebo plus vemurafenib, corresponding to a treatment difference of 4.68 points (95% CI, −2.05 to 11.40; P =0.17).

### NLME Modeling Characterized Longitudinal Treatment Effect

A NLME model was developed to characterize longitudinal trajectories in patient-reported GHS/QoL scores. The underlying PRO trajectories were represented by a pharmacology-informed nonlinear function, in which *Emax* described the magnitude of GHS/QoL change and *K_d_*described the rate of GHS/QoL change (**Figure 2a**). The model quantified within-patient variability, helping distinguish PRO changes related to disease and treatment from unexplained fluctuations that may reflect non-clinical factors (**Figure 2b**). The model also estimated the average trajectory across the study population while allowing each participant to have a unique trajectory, thereby capturing between-patient variability (**Figure 2c**). The final NLME model adequately reproduced the observed longitudinal patterns for both EORTC QLQ-C30 question 29 (overall health) and question 30 (overall quality of life) (**Figure 2d**). Additional model diagnostic results and final parameter estimates are provided in **Figure S1** and **Tables S2** and **S3**, respectively.

**Figure 2.**
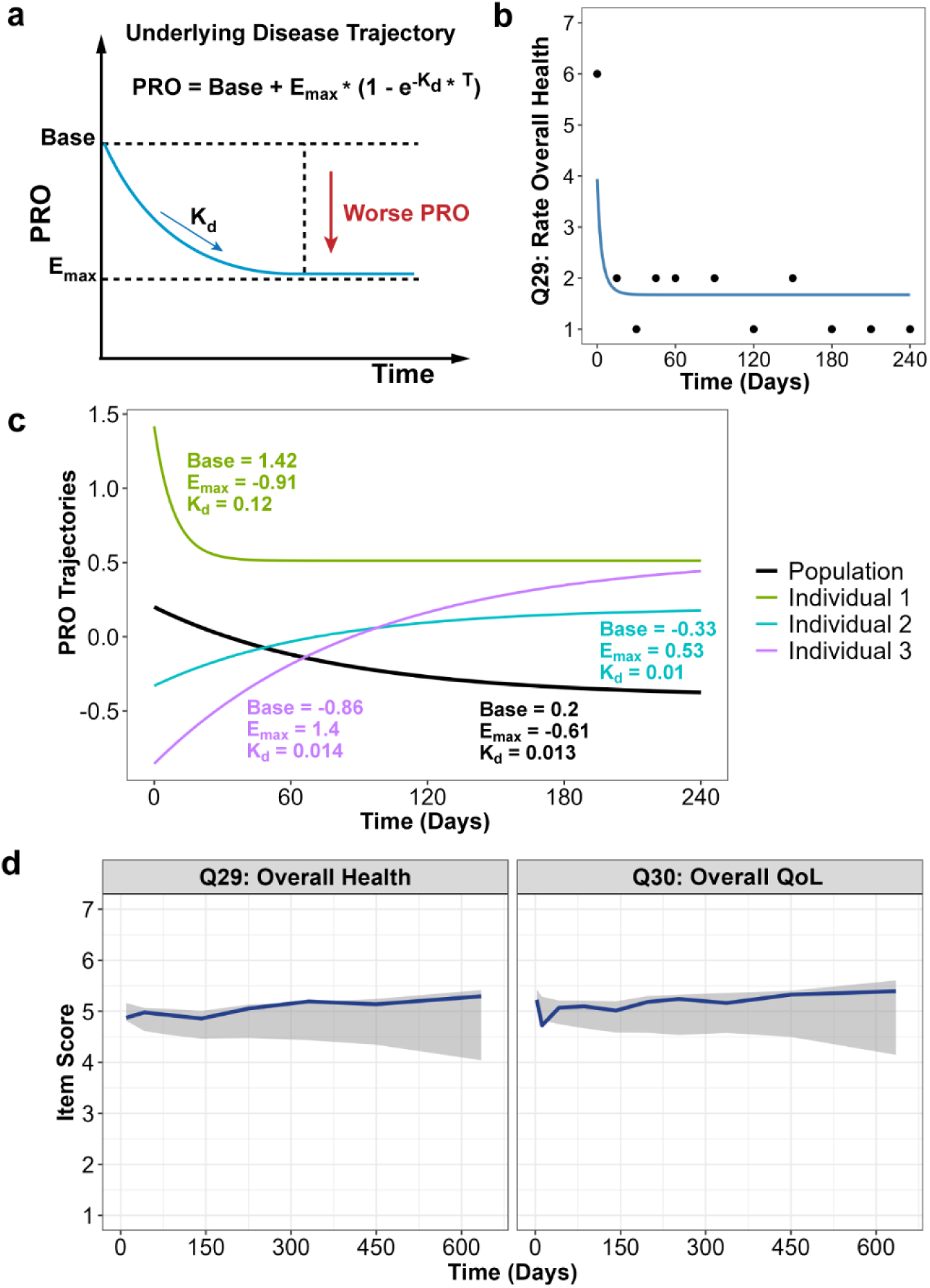
NLME modeling of longitudinal PRO trajectories. **(a)** Schematic representation of the nonlinear longitudinal PRO model. *Base* represents the baseline latent PRO variable, *Emax* represents the magnitude of GHS/QoL change, and *K_d_*describes the rate of PRO change. **(b)** Representative individual profile illustrating the model-estimated PRO trajectory (blue line) and within-patient variability in observed PRO score (black dots). **(c)** Population-average (black line) and representative individual PRO trajectories illustrating between-patient variability. **(d)** Visual predictive check (VPC) for EORTC QLQ-C30 question 29 and question 30. Solid lines represent the median observed items scores over time for each question, and shaded areas indicate the 95% model-predicted intervals.

After accounting for between-patient and within-patient variability, the NLME model provided an estimation of differences in longitudinal PRO trajectories between treatment groups. GHS/QoL remained relatively stable over time with cobimetinib plus vemurafenib but declined with placebo plus vemurafenib. The model-estimated PRO trajectory parameter *Emax* was significantly lower among participants receiving placebo plus vemurafenib than those receiving cobimetinib plus vemurafenib, indicating greater deterioration in GHS/QoL (P = 0.036, **Figure 3a**). After excluding the within-patient variability, the NLME model-estimated GHS/QoL scores showed smaller SDs (**Figure 2b**) than the observed GHS/QoL scores (**Figure 1**). Model-estimated GHS/QoL scores were significantly higher in cobimetinib plus vemurafenib arm than placebo plus vemurafenib at Cycle 8 Day 1(P < 0.05, **Figure 3b**).

**Figure 3.**
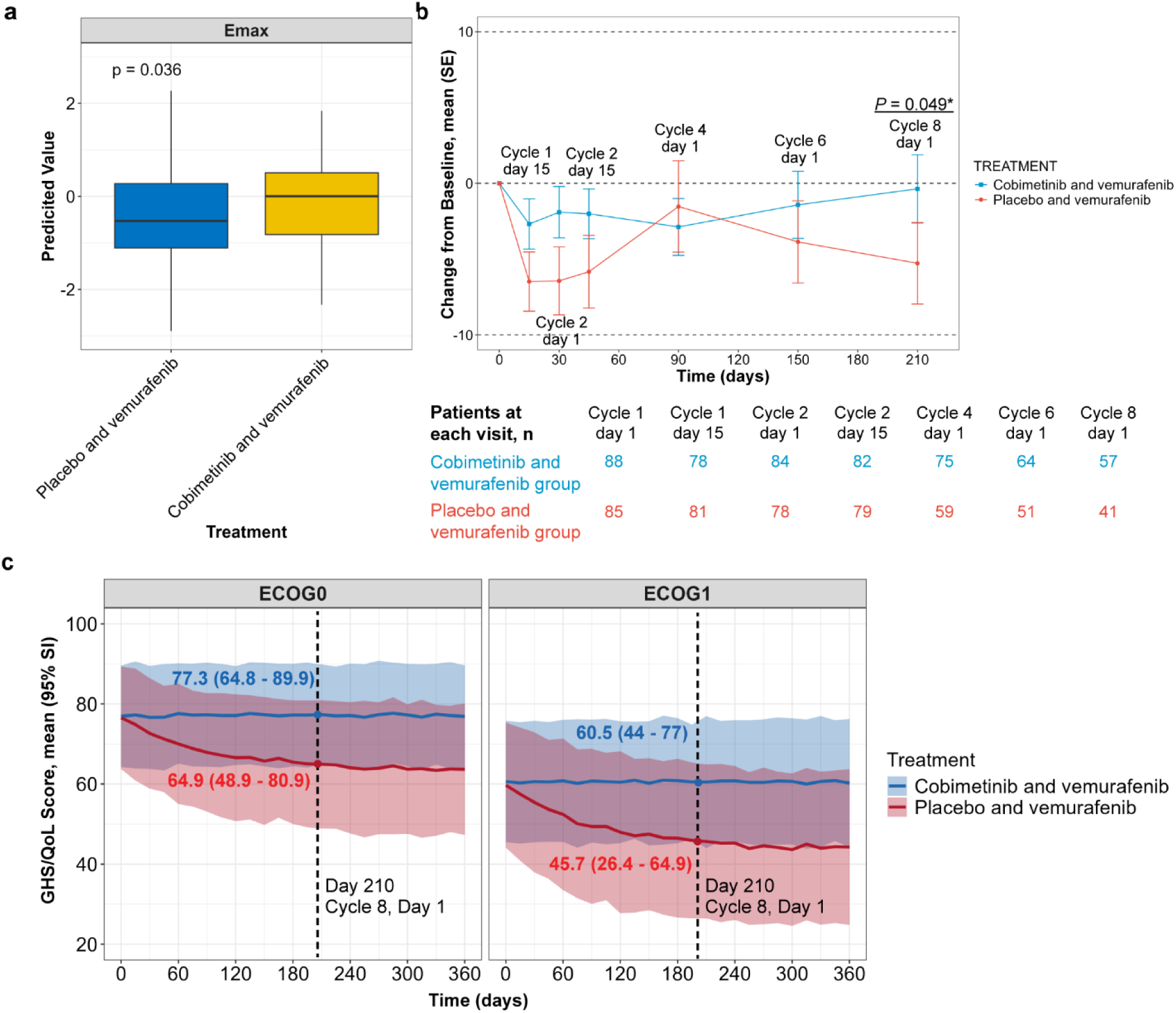
NLME model-based assessment of treatment effects on global health status/quality of life (GHS/QoL). **(a)** Distribution of individual model-estimated *E*max values by treatment arm. Lower *E*max values indicate greater deterioration in GHS/QoL. **(b)** Model-estimated mean changes from baseline in GHS/QoL over time by treatment arm. The dashed horizontal lines at ±10 points indicate the commonly used threshold for clinically meaningful improvement or deterioration. The numbers of evaluable patients at each visit are shown below the panel. **(c)** Model-simulated GHS/QoL trajectories for a typical virtual patient receiving cobimetinib plus vemurafenib or placebo plus vemurafenib, stratified by Eastern Cooperative Oncology Group (ECOG) performance status (0 or 1). Solid lines represent mean simulated trajectories and shaded areas represent 95% simulation intervals. The vertical dashed line indicates Cycle 8 Day 1 (day 210).

The treatment benefit was also clinically meaningful in NLME model simulations of a representative patient, accounting for model uncertainty (**Figure 3c**). At Cycle 8 Day 1, model-simulated GHS/QoL scores were better in cobimetinib plus vemurafenib than placebo plus vemurafenib for a representative patient with ECOG performance status 0 (77.3 [95% Simulation Interval (SI), 64.8-89.9] vs 64.9 [95% SI, 48.9-80.9]). Similar benefits were observed for a representative patient with ECOG performance status 1 (60.5 [95% SI, 44.0-77.0] vs 45.7 [95% SI, 26.4-64.9]). At Cycle 8 Day 1, model-based simulations predicted a difference in GHS/QoL favoring cobimetinib plus vemurafenib, with a mean between-group difference of 10.4 points (95% SI, -6.2, 26.9), exceeding the commonly used 10-point clinically meaningful threshold. [28]

### Association between PROs and Overall Survival

In Cox proportional hazards analyses, NLME model-estimated PRO trajectory parameters showed directional associations with overall survival, although these associations did not reach statistical significance (p> 0.05, **Figure S2**). Greater model-estimated change in GHS/QoL over time (*Emax*) was associated with worse overall survival (Hazard ratio 0.92 with 95% CI [0.74 – 1.15]), whereas higher baseline GHS/QoL was associated with better overall survival (Hazard ratio 0.82 with 95% CI [0.65 – 1.04]). These findings suggest that longitudinal PRO trajectories may contain prognostic information related to survival.

## Discussion

In this secondary analysis of the phase 3 coBRIM trial, NLME modeling identified a treatment-related difference in longitudinal GHS/QoL that was not apparent from either the observed scores or MMRM analysis. Observed GHS/QoL scores showed substantial variability, and the between-group difference at Cycle 8 Day 1 was not statistically significant. After accounting for between-patient and within-patient variability, the NLME model estimated a smaller GHS/QoL deterioration with cobimetinib plus vemurafenib than with placebo plus vemurafenib. Model-predicted GHS/QoL scores were also significantly higher with combination treatment at Cycle 8 Day 1, with an estimated between-group difference exceeding the commonly used 10-point clinically meaningful threshold. These findings suggest that NLME modeling may provide complementary information about treatment-related changes in PROs when substantial variability makes differences in observed PRO scores difficult to detect.

PRO data are inherently variable, and substantial variability can obscure treatment-related changes and complicate the interpretation of treatment effects [9]. NLME modeling addresses this challenge through two key advantages. First, it distinguishes underlying longitudinal PRO changes from between-patient and within-patient variability, allowing more precise characterization of treatment-related trajectories [26]. Second, it incorporates pharmacology-informed nonlinear functions to describe the magnitude and time course of PRO changes, which may reflect the combined influences of disease progression, therapeutic efficacy, and treatment-related toxicity. [19; 27] In our study, NLME model-estimated GHS/QoL scores showed smaller SDs than observed scores and revealed treatment-related differences that were not statistically significant in conventional analyses. These findings suggest that NLME modeling can provide complementary insights into longitudinal treatment effects that may be overlooked when analyzing highly variable observed PRO scores.

Our findings have important clinical implications for evaluating treatment benefit in oncology trials. Survival and tumor-based outcomes remain central measures of efficacy, but they do not fully describe how patients feel and function during treatment [2; 29]. PROs provide this complementary perspective and are increasingly incorporated into oncology drug development and regulatory evaluation. However, their value depends on the ability to interpret longitudinal changes in the presence of substantial variability. Our approach addresses important limitations of conventional hypothesis testing for PROs, which typically focuses on treatment differences at prespecified time points and may not fully capture the longitudinal trajectory and heterogeneity of patient-reported health status over the course of treatment. By modeling longitudinal PROs over time, our approach provides a more comprehensive characterization of treatment-related changes and can identify clinically meaningful patterns of response that may be overlooked by analyses based on a single endpoint or time point. This longitudinal perspective may provide clinicians and other decision-makers with more informative evidence regarding the magnitude, timing, and persistence of treatment effects, thereby supporting a more comprehensive evaluation of the benefits of cancer therapies and informing treatment decisions.

Several limitations should also be considered. This was a retrospective secondary analysis of one clinical trial, and the findings require validation across other treatments, cancer types, and PRO instruments. In addition, participant-level data were available for only 202 of the 495 participants enrolled in the parent coBRIM trial due to sponsor data restrictions, and the analytic cohort may therefore not fully represent the entire trial population. The subset of participants available was determined by sponsor and was not selected by the investigators based on PRO availability or clinical outcomes. In addition, the GHS/QoL trajectories simulated for representative patients were generated from the fitted model and should not be considered as additional independent clinical observations. Nevertheless, these findings demonstrate the potential for NLME modeling to complement conventional PRO analyses by better characterizing longitudinal treatment effects in highly variable patient-reported data.

In conclusion, pharmacology-informed NLME modeling identified a clinically meaningful longitudinal GHS/QoL treatment effect that was not evident from observed-score or MMRM analyses. By accounting for between-patient and within-patient variability and longitudinal PROs over time, this approach may complement conventional PRO analyses and improve characterization of treatment-related changes in patient-reported health status. Our study provides proof-of-concept for the broader application of NLME modeling to strengthen the interpretation and utilization of PRO evidence in oncology clinical trials.

## Data Availability

The source data used in this study were provided through the Vivli data-sharing platform. Access to the data is subject to Vivli data access procedures and applicable data use agreements.

## Acknowledgments

This publication is based on research using data from data contributor Roche that has been made available through Vivli, Inc. Vivli has not contributed to or approved, and is not in any way responsible for, the contents of this publication. The authors acknowledge the use of ChatGPT (OpenAI) for assistance with language editing and improving the clarity and readability of the manuscript. The authors reviewed and revised all AI-assisted text and take full responsibility for the final content.

## References

1. Siegel, R. L., Kratzer, T. B., Wagle, N. S., Sung, H., & Jemal, A. (2026). Cancer statistics, 2026. CA Cancer J Clin, 76(1), e70043.

2. Basch, E. (2016). Missing Patients’ Symptoms in Cancer Care Delivery--The Importance of Patient-Reported Outcomes. JAMA Oncol, 2(4), 433–434.

3. Basch, E. (2017). Patient-Reported Outcomes - Harnessing Patients’ Voices to Improve Clinical Care. N Engl J Med, 376(2), 105–108.

4. US Food and Drug Administration. Core patient-reported outcomes in cancer clinical trials: guidance for industry. (2024). Oncology Center of Excellence, Center for Biologics Evaluation and Research, and Center for Drug Evaluation and Research.

5. Kluetz, P. G., Slagle, A., Papadopoulos, E. J., Johnson, L. L., Donoghue, M., Kwitkowski, V. E., Chen, W. H., Sridhara, R., Farrell, A. T., Keegan, P., Kim, G., & Pazdur, R. (2016). Focusing on Core Patient-Reported Outcomes in Cancer Clinical Trials: Symptomatic Adverse Events, Physical Function, and Disease-Related Symptoms. Clin Cancer Res, 22(7), 1553–1558.

6. Blackstone, E. C., Efficace, F., Meyerhardt, J. A., & Abel, G. A. (2025). Timely Reporting of Patient-Reported Outcomes in Cancer Clinical Trials: An Ethical Imperative. J Clin Oncol, 43(10), 1176–1179.

7. Basu, A., & Hershman, D. L. (2024). The “PRO” mise and “PRO” gress of PROs in cancer clinical trials (Vol. 116, pp. 1544–1546): Oxford University Press.

8. Shahraz, S., Wong, O. Y., Chotai, R., Knight, S., Globe, D., Patel, M. J., Kaushik, A., Aiyegbusi, O. L., & Calvert, M. (2026). Communicating clinical trial information about cancer drug products to patients and healthcare professionals: review of patient-reported outcome messaging. BMJ Oncol, 5(1), e000942.

9. Zhou, J., Muluneh, B., Li, Q., & Hughes, J. H. (2025). Revolutionizing Patient-Reported Outcomes Analysis for Oncology Drug Development Using Population Models. Clin Cancer Res, 31(9), 1580–1586.

10. Miaskowski, C., Dunn, L., Ritchie, C., Paul, S. M., Cooper, B., Aouizerat, B. E., Alexander, K., Skerman, H., & Yates, P. (2015). Latent Class Analysis Reveals Distinct Subgroups of Patients Based on Symptom Occurrence and Demographic and Clinical Characteristics. J Pain Symptom Manage, 50(1), 28–37.

11. Miaskowski, C., Cooper, B. A., Melisko, M., Chen, L. M., Mastick, J., West, C., Paul, S. M., Dunn, L. B., Schmidt, B. L., Hammer, M., Cartwright, F., Wright, F., Langford, D. J., Lee, K., & Aouizerat, B. E. (2014). Disease and treatment characteristics do not predict symptom occurrence profiles in oncology outpatients receiving chemotherapy. Cancer, 120(15), 2371–2378.

12. Zhou, J., Muluneh, B., Wang, Z., Yao, H., & Hughes, J. H. (2025). Leveraging Longitudinal Patient-Reported Outcome Trajectories to Predict Survival in Non-Small Cell Lung Cancer. Clin Cancer Res, 31(13), 2685–2694.

13. Stone, A. A., Broderick, J. E., Schneider, S., & Schwartz, J. E. (2012). Expanding options for developing outcome measures from momentary assessment data. Psychosom Med, 74(4), 387–397.

14. Lee, M. K., Basch, E., Mitchell, S. A., Minasian, L. M., Langlais, B. T., Thanarajasingam, G., Ginos, B. F., Rogak, L. J., Mendoza, T. R., Bennett, A. V., Schrag, D., Mazza, G. L., & Dueck, A. C. (2023). Reliability and validity of PRO-CTCAE® daily reporting with a 24-hour recall period. Qual Life Res, 32(7), 2047–2058.

15. Jim, H. S., Small, B., Faul, L. A., Franzen, J., Apte, S., & Jacobsen, P. B. (2011). Fatigue, depression, sleep, and activity during chemotherapy: daily and intraday variation and relationships among symptom changes. Ann Behav Med, 42(3), 321–333.

16. Sun, Y., Chang, S., Tang, K., LeBlanc, M. R., Palmer, A. C., Ahamadi, M., & Zhou, J. (2026). When survival improves but quality of life does not: a model-based meta-analysis of immune checkpoint inhibitors. J Natl Cancer Inst.

17. Shahnam, A., Nindra, U., Desai, J., Hui, R., Buyse, M., Hopkins, A. M., & Sorich, M. J. (2023). Time to deterioration of patient-reported outcomes as a surrogate of overall survival: a meta-analysis. J Natl Cancer Inst, 115(12), 1475–1482.

18. Sherry, A. D., Miller, A. M., Parlapalli, J. P., Kupferman, G. S., Beck, E. J., McDonald, J., Kouzy, R., Abi Jaoude, J., Lin, T. A., Sanford, N. N., Chino, F., Gyawali, B., Booth, C., Msaouel, P., & Ludmir, E. B. (2025). Overall Survival and Quality-of-Life Superiority in Modern Phase 3 Oncology Trials: A Meta-Epidemiological Analysis. JAMA Oncol, 11(7), 718–724.

19. Mould, D. R., & Upton, R. N. (2012). Basic concepts in population modeling, simulation, and model-based drug development. CPT Pharmacometrics Syst Pharmacol, 1(9), e6.

20. Llanos-Paez, C., Ambery, C., Yang, S., Beerahee, M., Plan, E. L., & Karlsson, M. O. (2022). Improved Confidence in a Confirmatory Stage by Application of Item-Based Pharmacometrics Model: Illustration with a Phase III Active Comparator-Controlled Trial in COPD Patients. Pharm Res, 39(8), 1779–1787.

21. Ascierto, P. A., McArthur, G. A., Dréno, B., Atkinson, V., Liszkay, G., Di Giacomo, A. M., Mandalà, M., Demidov, L., Stroyakovskiy, D., Thomas, L., de la Cruz-Merino, L., Dutriaux, C., Garbe, C., Yan, Y., Wongchenko, M., Chang, I., Hsu, J. J., Koralek, D. O., Rooney, I., Ribas, A., & Larkin, J. (2016). Cobimetinib combined with vemurafenib in advanced BRAF(V600)-mutant melanoma (coBRIM): updated efficacy results from a randomised, double-blind, phase 3 trial. Lancet Oncol, 17(9), 1248–1260.

22. Dréno, B., Ascierto, P. A., Atkinson, V., Liszkay, G., Maio, M., Mandalà, M., Demidov, L., Stroyakovskiy, D., Thomas, L., de la Cruz-Merino, L., Dutriaux, C., Garbe, C., Bartley, K., Karagiannis, T., Chang, I., Rooney, I., Koralek, D. O., Larkin, J., McArthur, G. A., & Ribas, A. (2018). Health-related quality of life impact of cobimetinib in combination with vemurafenib in patients with advanced or metastatic BRAF(V600) mutation-positive melanoma. Br J Cancer, 118(6), 777–784.

23. Rosen, L. S., LoRusso, P., Ma, W. W., Goldman, J. W., Weise, A., Colevas, A. D., Adjei, A., Yazji, S., Shen, A., Johnston, S., Hsieh, H. J., Chan, I. T., & Sikic, B. I. (2016). A first-in-human phase I study to evaluate the MEK1/2 inhibitor, cobimetinib, administered daily in patients with advanced solid tumors. Invest New Drugs, 34(5), 604–613.

24. Fayers, P., & Bottomley, A. (2002). Quality of life research within the EORTC-the EORTC QLQ-C30. European Organisation for Research and Treatment of Cancer. Eur J Cancer, 38 Suppl 4, S125–133.

25. Aaronson, N. K., Ahmedzai, S., Bergman, B., Bullinger, M., Cull, A., Duez, N. J., Filiberti, A., Flechtner, H., Fleishman, S. B., de Haes, J. C., &, et al. (1993). The European Organization for Research and Treatment of Cancer QLQ-C30: a quality-of-life instrument for use in international clinical trials in oncology. J Natl Cancer Inst, 85(5), 365–376.

26. Mehta, K., Hughes, J. H., Zhou, J., Muluneh, B., & Gnanasakthy, A. (2026). Tutorial on Analyzing Patient-Reported Outcome Measures in Clinical Development. Clin Transl Sci, 19(8), e70675.

27. Holford, N. (2017). Pharmacodynamic principles and the time course of immediate drug effects. Transl Clin Pharmacol, 25(4), 157–161.

28. Cocks, K., King, M. T., Velikova, G., Martyn St-James, M., Fayers, P. M., & Brown, J. M. (2011). Evidence-based guidelines for determination of sample size and interpretation of the European Organisation for the Research and Treatment of Cancer Quality of Life Questionnaire Core 30. J Clin Oncol, 29(1), 89–96.

29. Bouchard, L. C., Aaronson, N., Gondek, K., & Cella, D. (2018). Cancer symptom response as an oncology clinical trial end point. Expert Rev Qual Life Cancer Care, 3(2-3), 35–46.

