## Supplementary Materials for "Nonlinear Mixed Effect Modeling of Patient-Reported Outcomes to Improve Treatment Effect Estimation in an Oncology Clinical Trial"

### Supplementary Methods

#### 1. Item-response theory (IRT) model for individual symptoms

The IRT model was developed in two steps: (1) a base IRT model was constructed to link the probability of each observed response to an underlying latent quality-of-life (QoL) variable; (2) a longitudinal structural model was built to describe the trajectory of this latent variable over time.

*Base IRT Model*

The IRT describes the probability of each response category as a function of patients' underlying QoL latent variable ($L$), representing patient status [1, 2].

Our IRT model applied the same latent variable to both Question 29 and Question 30 from EORTC QLQ-C30. The graded response model was used to characterize each question [1, 3], where probability for subject $i$ to have at least a response of $s$ (s = 1 to 7) for question $j$ was described in the equation 1 and the probability of having the score s is calculated based on equation 2.

$$P(y_{ij} \geq s) = \frac{1}{1+e^{-a(L_i - b_{j,s})}} \quad \text{(Equation 1)}$$

$$P(y_{ij} = s) = P(y_{ij} \geq s) - P(y_{ij} \geq s+1) \quad \text{(Equation 2)}$$

where $a$ is the discrimination parameter shared by Q29 and Q30, and $b_{j,s}$is the difficulty (threshold) parameter for response level s of question $j$. The difficulty parameters were constrained to be non-decreasing within each item $(b_{j,1} < b_{j,3} < \cdots < b_{j,7})$. $L_i$ represents the latent global quality-of-life variable for patient $i$, which is shared by both Q29 and Q30. A higher $L_i$value indicates a better perceived global health status and quality of life. $L_i$ was assumed to follow a standard normal distribution, $L_i(0) \sim N(0,1)$, at baseline and estimated in the later longitudinal model. Item characteristic curves (ICC) of each question were generated to show the probability of each response level for a given level of patient QoL latent variable ($L$). The shape of the ICCs is determined by $a$ and $b_{j,s}$.

*Longitudinal IRT Model*

An asymptotic equation was used to characterize the longitudinal changes of the QoL status of each subject:

$$L_i(t) = L_{i,BASE} + E_{\max_i} \times \left(1 - e^{-K_{d_i} t}\right) \qquad \text{(Equation 3)}$$

where $L_i(t)$ is latent QoL status at time *t* in days for subject *i* and $L_{i,BASE}$ is the latent QoL status at baseline for subject *i*. $E_{\max_i}$ is the maximum latent QoL improvement for subject *i*. $K_{d_i}$ is the latent QoL improvement rate for subject *i*.

**1.3 NLME model development**

To explain the heterogeneity across subjects, inter-individual variability (IIV) was incorporated into the NLME model parameters. The IIV was described using a normal or log-normal distribution, as defined in Equations (4) and (5) respectively

$P_i = \theta_P + \eta_i$ (Equation 4)

$P_i = \theta_P \cdot e^{\eta_i}$ (Equation 5)

Where $P_i$ represents the individual parameter value for subject $i$, $\theta_P$ is the typical population value of parameter $P$, and $\eta_i$ is a random effect assumed to follow a normal distribution with mean 0 and variance $\omega^2$. IIV was assumed to be a normal distribution for parameter $L_{BASE}$, $E_{max}$ and a log-normal distribution for parameter $K_d$.

**1.4 Covariate model development**

A covariate model was developed to evaluate the effect of the potential baseline covariates on relevant model parameters based on exploratory analysis and clinical relevance. Investigated baseline covariates included patient demographics (sex, age, race and ethnicity), and Eastern Cooperative Oncology Group (ECOG) performance status. Covariate analysis was performed using the stepwise covariate model building procedure (SCM), with a significance level of $p < 0.01$ in the forward selection and $p < 0.001$ in the backward elimination. The effect of a categorical covariate on a parameter was described as a linear relationship as shown in Equation (6) and (7). Centered linear relationships were used to describe the effect of a continuous covariate on a parameter as shown in equation (8).

$$P_i = \theta_P + \theta_{COV} \quad \text{(Equation 6)}$$

$$\theta_{COV} = \begin{cases} 0 & if\ reference\ category \\ \theta_{COV,1} & if\ category\ 1 \\ \theta_{COV,2} & if\ category\ 2 \\ & \cdots \end{cases} \quad \text{(Equation 7)}$$

$$P_i = \theta_P + \theta_{COV}(COV_i - COV_{median}) \quad \text{(Equation 8)}$$

where $P_i$ is the individual parameter value, $\theta_P$ is the typical value of the parameter $P$, $\theta_{COV}$ is the coefficient for the effect of the covariate on parameter $P$. $COV_i$ is the continuous covariate value for subject $i$ and $COV_{median}$ is the median value of the covariate in the population. Missing values for covariates were imputed with the population median (for continuous covariates) or mode (for categorical covariates) during the covariate analysis.

**1.5 Model selection**

Model selection was based on the minimum objective function value (OFV), diagnostic plots and the precision of parameter estimations. For nested models, a decrease in OFV of ≥ 6.63 ($p < 0.01$) was considered a statistically significant improvement. The predictive performance of the models was assessed through visual predictive checks (VPCs). VPC is a tool in NLME models to compare observed data to model-simulated prediction intervals to assess the model's ability to capture the central tendency and variability of the data. [4, 5] A total of 500 Monte Carlo simulations of the dataset were performed to generate the VPCs. Parameter estimates were assessed by two metrics: (1) Relative standard error (RSE) reflects the precision of parameter estimates. Generally, RSE values below 40% indicate stable and reliable parameter estimation. (2) Shrinkage is the parameter indicating how much individual parameter estimates were pulled towards population typical parameter values. Shrinkage < 30% suggests the model captures sufficient between-subject variability. A high shrinkage (> 60%) means the individual predictions may not be reliable—even when model diagnostics look acceptable overall.

**References**


1. Schindler E, Friberg LE, Lum BL*, et al.* A Pharmacometric Analysis of Patient-Reported Outcomes in Breast Cancer Patients Through Item Response Theory. Pharm Res 2018;35(6):122.

2. Van der Linden WJ, Hambleton RK. *Handbook of modern item response theory*: Springer Science & Business Media; 2013.

3. Ueckert S. Modeling Composite Assessment Data Using Item Response Theory. CPT Pharmacometrics Syst Pharmacol 2018;7(4):205-218.

4. Bergstrand M, Hooker AC, Wallin JE*, et al.* Prediction-corrected visual predictive checks for diagnosing nonlinear mixed-effects models. Aaps j 2011;13(2):143-51.

**Supplementary Figures**

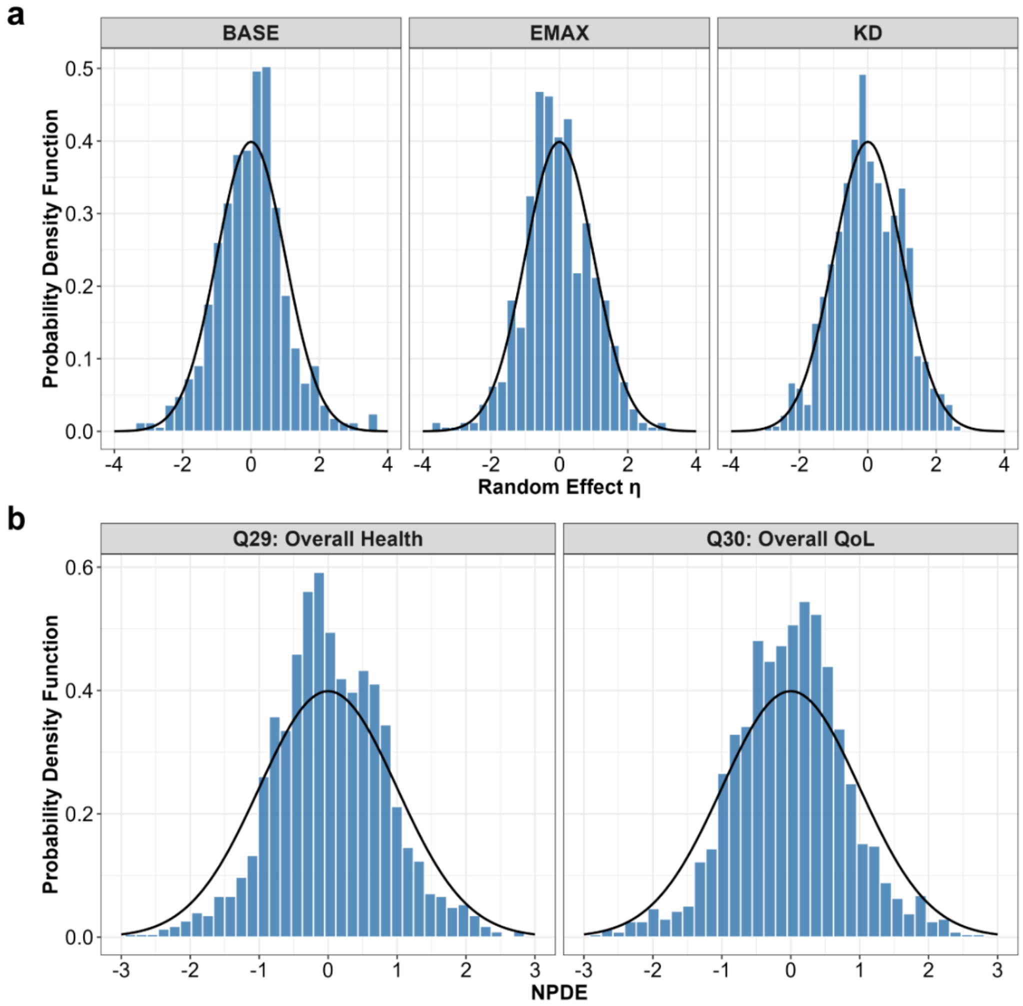


**Figure S1: Diagnostic plots for the final longitudinal IRT model.**

(a) Distributions of individual random effects for Base, $E_{max}$, and $K_D$; (b) Normalized prediction distribution error (NPDE) distributions for Q29 overall health and Q30 overall quality of life. NPDE, normalized prediction distribution error; QoL, quality of life.

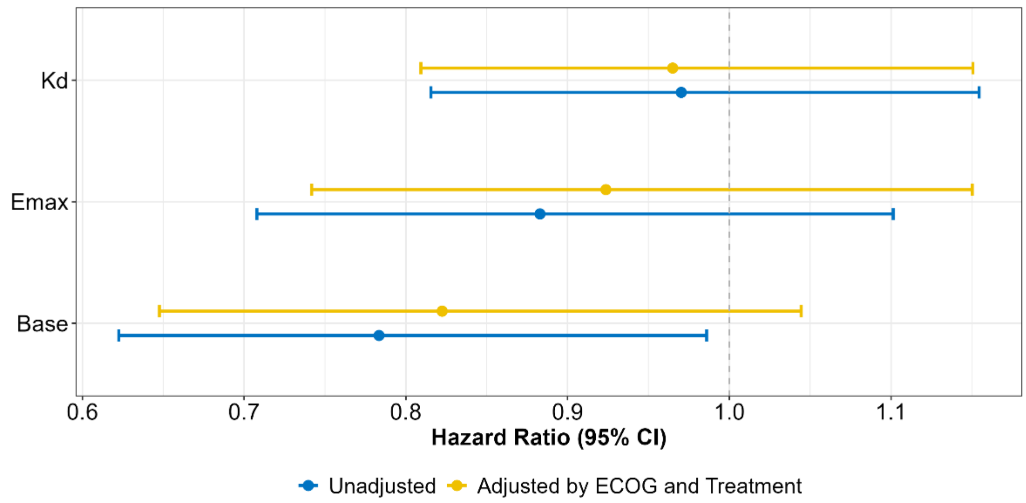


**Figure S2: Association between individual IRT model parameters and overall survival.** Hazard ratios (HRs) and 95% confidence intervals (CIs) were estimated using multivariable Cox proportional hazards regression models. Blue estimates represent the model without additional clinical covariates, whereas yellow estimates represent the model stratified by Eastern Cooperative Oncology Group (ECOG) performance status and adjusted for treatment arms.

**Supplementary Tables**

**Table S1. MMRM-Estimated Least-Squares Mean Changes From Baseline in Quality-of-Life Scores and Treatment Differences by Visit**

| Visit | Active<br>Mean (SE) | Control<br>Mean (SE) | Difference<br>Effect, 95% CI | p-value |
|---|---|---|---|---|
| Cycle 1 Day 15 | -8.63 (2.49) | -11.47 (2.45) | 2.84 (-4.07, 9.74) | 0.42 |
| Cycle 2 Day1 | -3.58 (1.94) | -2.63 (1.98) | -0.95 (-6.43, 4.54) | 0.73 |
| Cycle 2 Day15 | -0.42 (2.0) | -4.92 (2.01) | 4.51 (-1.1, 10.11) | 0.11 |
| Cycle 4 Day1 | -3.95 (2.17) | -2.03 (2.34) | -1.92 (-8.22, 4.38) | 0.55 |
| Cycle 6 Day1 | -2.51 (2.08) | -4.44 (2.25) | 1.93 (-4.14, 8.0) | 0.53 |
| Cycle 8 Day1 | -2.84 (2.23) | -7.52 (2.55) | 4.68 (-2.05, 11.4) | 0.17 |

LS mean difference calculated as Treatment − Placebo.

**Table S2: Item-specific parameter estimates from the final longitudinal item-response theory model.**

| **Item** | a | b1 | $\delta_2$ | $\delta_3$ | $\delta_4$ | $\delta_5$ | $\delta_6$ |
|---|---|---|---|---|---|---|---|
| Q29: Overall Health | 4.94 | -2.9 | 1.04 | 0.7 | 0.58 | 0.75 | 1.1 |
| Q30: Overall Quality of Life | 4.94 | -2.74 | 0.87 | 0.65 | 0.52 | 0.72 | 0.87 |

$b_2 = b_1 + \delta_2$; $b_3 = b_2 + \delta_3$; $b_4 = b_3 + \delta_4$; $b_5 = b_4 + \delta_5$; $b_6 = b_5 + \delta_6$

**Table S3. Final Item Response Theory model parameter estimates**

| **Parameter** | Value | RSE (%) | SHR (%) |
|---|---|---|---|
| **Population Parameters** | | | |
| Baseline GHS/QoL Latent Variable (Base) | 0.2 | 38.2 | 4.56 |
| Maximum change of GHS/QoL Latent Variable ($E_{max}$) | 0 fixed | NA | 23.9 |
| GHS/QoL Latent Variable Change Rate ($K_d$, 1/day) | 0.013 | 47.1 | 58.4 |
| **Inter-individual Variability** | | | |
| Standard Deviation of Base ($\Omega_{BSV\_Base}$) | 0.86 | 5.71 | |
| Standard Deviation of $E_{max}$($\Omega_{BSV\_E_{max}}$) | 1.54 | 8.56 | |
| Standard Deviation of $K_d$ ($\Omega_{BSV\_K_D}$ ) | 3.51 | 12.0 | |
| **Covariates** | | | |
| ECOG I on Base | -0.73 | 18.7 | |
| Placebo Arm on $E_{max}$ | -0.61 | 31.7 | |

GHS/QoL, Global health status/Quality of Life. RSE, relative standard error. SHR, shrinkage.
